# Identifiability of infection model parameters early in an epidemic

**DOI:** 10.1101/2020.06.15.20132217

**Authors:** Timothy Sauer, Tyrus Berry, Donald Ebeigbe, Michael M. Norton, Andrew Whalen, Steven J. Schiff

## Abstract

It is known that the parameters in the deterministic and stochastic SEIR epidemic models are structurally identifiable. For example, from knowledge of the infected population time series *I*(*t*) during the entire epidemic, the parameters can be successfully estimated. In this article we observe that estimation will fail in practice if only infected case data during the early part of the epidemic (pre-peak) is available. This fact can be explained using a long-known phenomenon called dynamical compensation. We use this concept to derive an unidentifiability manifold in the parameter space of SEIR that consists of parameters indistinguishable to *I*(*t*) early in the epidemic. Thus, identifiability depends on the extent of the system trajectory that is available for observation. Although the existence of the unidentifiability manifold obstructs the ability to exactly determine the parameters, we suggest that it may be useful for uncertainty quantification purposes. A variant of SEIR recently proposed for COVID-19 modeling is also analyzed, and an analogous unidentifiability surface is derived.

## 1. Introduction

In nonlinear systems, identifiability of parameters depends critically on location in phase space. In this article, we point out a particularly vivid illustration of this fact that occurs in SEIR (Susceptible, Exposed, Infected, Removed) modeling of epidemics. While the SEIR parameters are identifiable from the infected population *I*(*t*) if the entire epidemic is observed, the ability to infer parameters from the pre-peak portion of the epidemic is strictly limited, due to the approximately linear dynamics that occur early in the epidemic.

We explain this failure of identifiability in Section 3, where we show that for a given instance of the infected time series *I*(*t*) early in the epidemic, there are multiple solutions with various parameters values that are approximately consistent with the same *I*(*t*). Moreover, we show that these multiple solutions form a two-dimensional *unidentifiability manifold* that indexes the alternative parameter sets that are consistent with *I*(*t*). The alternate parameter sets on this surface share the growth rate of the epidemic (the leading eigenvalue of the linearized system) even though their respective parameter values vary widely. Thus estimating all parameters solely from knowledge of the infected cases during the pre-peak portion of the trajectory is not possible in practice, with any parameter estimation algorithm.

Since the unidentifiability set is two-dimensional, it follows that two of the three unknown parameters (in the basic SEIR) must be known *a priori* in order to determine the third. In particular, the reproductive number *R*_0_, which is often derived from two of the SEIR parameters, is in practice not identifiable from *I*(*t*) alone.

Unidentifiability is an underappreciated issue in viral modeling. The authors of the comprehensive review [12] state that mathematical modeling of epidemics “usually overparameterizes the model and ignores parameter identifiability, which makes it difficult to directly fit such models to data.” We corroborate this opinion by showing that it is impossible in practice to determine more than one unknown SEIR parameter from observations of *I*(*t*) preceding the peak stage of the epidemic, and exhibit the underlying mathematical reasons. While overparametrization is rampant in the literature, our focus here is deliberately on a reasonably-parametrized epidemic model, which suffers from unidentifiability only in a crucial region of phase space.

We will refer to this deficiency as *trajectory-dependent unidentifiability*. The difficulty stems from a phenomenon called *dynamical compensation* [18], as identified in linear compartmental models by Bellman and Aström [2] in 1970. In the terminology of [18], it is a *structural unidentifiability* [15, 19] in the linear model that approximates SEIR in the early portion of the epidemic, which gradually disappears as the non-linearities become significant as the epidemic progresses (see Figure 4). Determination of the full parameter set is possible if *I*(*t*) can be observed through the peak of the infection.

To illustrate identifiability issues that arise in applications, we employ two disparate approaches to parameter estimation. One is a parameter estimation algorithm based on data assimilation from partial observations, and the other an implementation of Markov Chain Monte Carlo (MCMC) techniques [8]. These are introduced in Section 2.2. These are two choices from several alternatives that are in common usage, some based directly on Bayesian inference [1], and others using data assimilation in more sophisticated ways [6, 9, 14]. The principal unidentifiability results of this article are independent of the method of parameter estimation applied.

Our analysis was preceded by work on dynamical compensation for linear systems, for example in [20], that shows how to find alternate parameter sets whose solutions do not change the observable *I*(*t*). These solutions are designed to match the true underlying solution even during the initial and often unobservable transient at the outset of the epidemic. However, by ignoring rapidly decaying dynamics at early times, our analysis uncovers a larger set of alternative parameters combinations that match observations. Somewhat counter-intuitively, it is exactly this expanded set of parameters that appear to be explored by parameter estimation methods, not the more restrictive parameter set [20]. This indicates that our simplifying assumptions allow us to correctly anticipate the performance of these methods (see Figure 5).

Despite the fact that the unidentifiability surface shows why exact determination of parameters is impossible during the pre-peak interval, it has a useful purpose for uncertainty quantification, because it constrains the set of alternative parameters that also generate *I*(*t*). Assume a parameter estimation algorithm is used to calculate a parameter set *p* from an observed *I*(*t*) early in the epidemic. Since the system is unidentifiable, another algorithm may provide another parameter set *p*′. However, we can expect it to lie on the unidentifiability surface of *p*, which is a constraint. We show in Section 4.3 that the systems corresponding to parameter sets chosen from the surface have dynamics much closer to the system generated by *p* than those chosen off the surface. By studying these nearby systems, we may be able to gain knowledge about the uncertainty of the system with estimated parameter set *p*′.

In Section 2 we review the deterministic and stochastic SEIR models and introduce two parameter estimation approaches. In Section 3 the notion of dynamical compensation is explored and its existence in a linearized version of SEIR is observed. The relevance to the problem of identifiability of parameters in the full nonlinear SEIR is noted in Section 4. In Section 5, the COVID-19 model of [14] is studied. A similar obstruction to identifiability caused by dynamical compensation is observed in this model.

## 2. Identifying parameters in SEIR

### 2.1. The deterministic and stochastic SEIR models

The deterministic version of the SEIR model [7, 11] that we will consider is

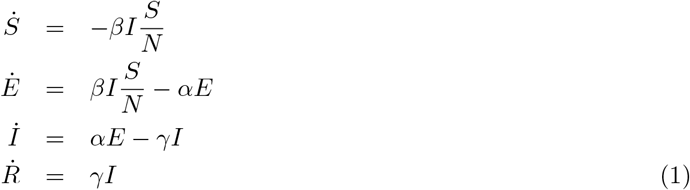

where the variables *S, E, I*, and *R* represent the populations of susceptible, exposed, infected, and removed patients, respectively and *N* = *S* + *E* + *I* + *R* denotes the total population. Time is measured in days. We use the simplest, or SEIR without vital statistics model, which assumes *N* to be constant with no births and deaths. There are more complex versions with additional parameters, but the identifiability issues we want to describe occur even for this simplest model. The sole nonlinearity is the *βIS/N* term which moves patients from the susceptible compartment to the exposed compartment according to transmission rate coefficient *β*.

We will interpret the model in the following way. The parameter *α* is the time constant of movement from exposed to infected; thus we assume that on average, the patient spends 1*/α* days as exposed before transitioning to infected, where we assume viral shedding begins. We will also make the assumption that symptoms are present in patients in the *I* compartment, so that the case can for the first time be observable. After 1*/γ* days in the *I* compartment, on average, the patient is removed from the population and does not return to the susceptible class.

Our principal interest is in determining what information can be inferred from measured reports of infected cases *I*(*t*). We address two obvious limitations of these assumptions. First, perhaps not all infected cases are reported. Thus, the true infected number may be *c*_1_*I* instead of *I*. Furthermore, a portion of the infected cases may be asymptomatic, and are not reported due to that reason. Thus, the true infected number may be *c*_2_*c*_1_*I*. In either case, the true number of infected may not be knowable. If the true number of infected is proportional to the reported *I*, the meaning of the contact transmission parameter *β* will be changed. However, many of the purposes of using the model, such as forecasts of future *I*(*t*), may still proceed unaffected.

In addition to the deterministic version, we will also consider the SEIR model as a set of stochastic differential equations with Poisson noise. In this version, we will calculate trajectories as follows. For each time step, the right-hand side of the equations will be evaluated by selecting from a Poisson distribution, and then integrated using an Euler method step. In other words, the values

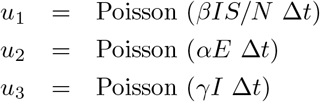

are chosen to represent the contribution of the right-hand side at each step, i.e.

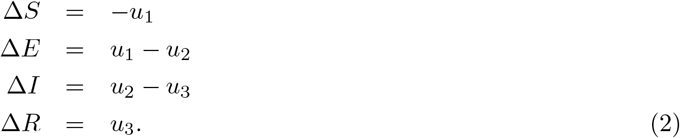

This version treats the SEIR model as a stochastic system for greater fidelity. However, our main conclusions about identifiability will be relevant for both the deterministic and stochastic versions.

### 2.2. Parameter estimation

Parameter estimation is customarily achieved by locating, implicitly or explicitly, the optimum of some auxiliary function that measures the fitness of the parameters. In some methods, the likelihood or marginal probability is maximized, while in others, an error or loss function is minimized.

In one method to estimate parameters *β, α*, and *γ* from daily reports of the single observable *I*(*t*), we will choose a particular loss function based on data assimilation, and explicitly minimize it. This approach will be useful to illustrate the geometry of the minima of the loss function in two different parts of the SEIR trajectory. Our choice for the loss function will be the data assimilation error in *I*(*t*) incurred while using the proposed set of parameters to optimally reconstruct the trajectory (*S*(*t*), *E*(*t*), *I*(*t*), *R*(*t*)) from the observed *I*(*t*). The use of data assimilation to reconstruct unobserved variables is the basis of modern numerical weather prediction, and has started to appear in epidemic modeling [4, 6, 9]. For the deterministic SEIR, we employ a standard Ensemble Kalman Filter (EnKF) [17, 16] to reconstruct the dynamics. For the stochastic SEIR, we use an EnKF tailored to Poisson noise instead of the standard Gaussian assumption. The EnKF used for this purpose is based on the Poisson Kalman Filter (PKF) from [5].

Data assimilation gives a way of reconstructing all variables of a differential equations model from partial observations, for example by measurements of one key variable. For SEIR model (1), if the parameters *β, α*, and *γ* are known, the observable *I*(*t*), or alternatively the daily changes Δ*I*(*t*) = *I*(*t*) − *I*(*t* −1), are in general sufficient to reconstruct the other three variables *S, E*, and *R*. Figure 1(a) shows a trajectory of a stochastic SEIR model (1) with parameters *β* = 1.1, *α* = 0.2, and *γ* = 0.5, and with initial conditions *S* = 10^6^, *E* = 10^2^, *I* = 0, *R* = 0. The inputs to the data assimilation algorithm are the model, the exact parameters, and the daily reports of new infections Δ*I*(*t*) = *I*(*t*) −*I*(*t*− 1). The assimilation algorithm uses the initial condition *S* = 10^6^, *E* = 0, *I* = 0, *R* = 0. That is, it is allowed to know the (constant) total population, but no information about the initial caseload. The EnKF is used to estimate the most likely values of *S*(*t*), *E*(*t*), *I*(*t*), and *R*(*t*) given the reports Δ*I*(*t*). Figure 1(b) shows the resulting reconstructed trajectory, a reasonably accurate version of the original.

**Figure 1:**
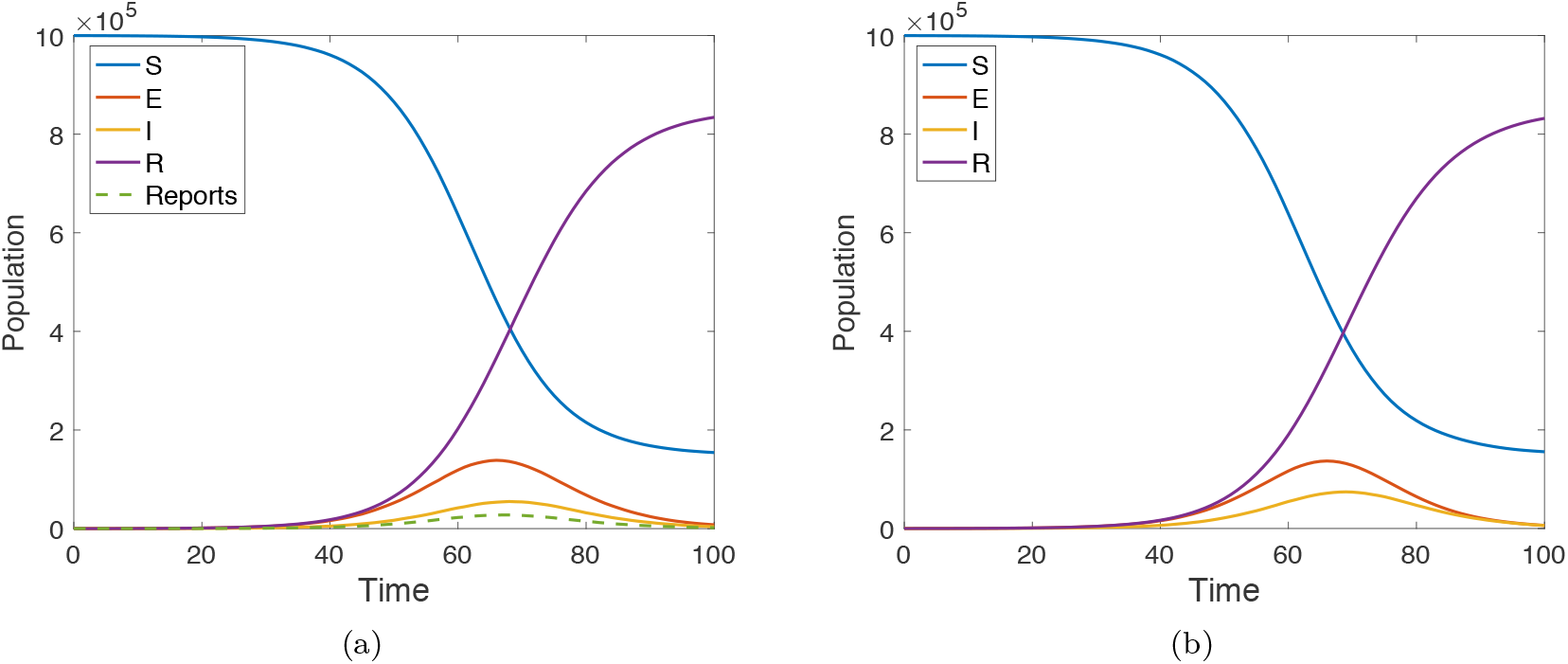
(a) Solution of the SEIR equations (1) with initial conditions *S* = 10^6^, *E* = 10^2^, *I* = 0, *R* = 0. The parameter settings are *β* = 1.1, *α* = 0.2, *γ* = 0.5. (b) Result of data assimilation using exact parameters of model with initial conditions *S* = 10^6^, *E* = 0, *I* = 0, *R* = 0, and the reports Δ*I* as inputs.

If the parameters are not known, and incorrect parameters are used in the model, the reconstruction in general will be farther from the original. This leads to a convenient loss function to consider for the purposes of parameter estimation. Let *L*(*β, α, γ*) denote the mean squared difference between the observed Δ*I*(*t*) and the reconstructed Δ*I*(*t*) from the EnKF, over a time interval [*T*_1_, *T*_2_]. Then minimization of *L* as a function of the parameters should lead to the correct, or generating, parameters.

To begin, we carried out this idea on the deterministic SEIR model (2) with a standard simplex minimization algorithm [13]. We started the simplex algorithm with 1000 starting guesses for the parameters *β, α, γ* that varied from the exact values by about 50%. Figure 2(a) shows the cumulative results of the minimization procedure for a trajectory of length 100 days, using two different intervals of observations, [*T*_1_, *T*_2_] = [0, 50] or [50, 100], with 1000 realizations of starting parameter guesses. There is a dramatic difference, depending on whether the time interval [0, 50] or [50, 100] is used for the input *I*(*t*). The red dottted curve is a histogram of approximate parameters using Δ*I*(*t*) from the interval [0, 50]. The black histogram uses the interval [50, 100]. While the histogram shows no identifiability on [0, 50], on the interval [50, 100] the method finds the correct parameters with less than 0.1% error on over 95% of the 1000 attempts.

**Figure 2:**
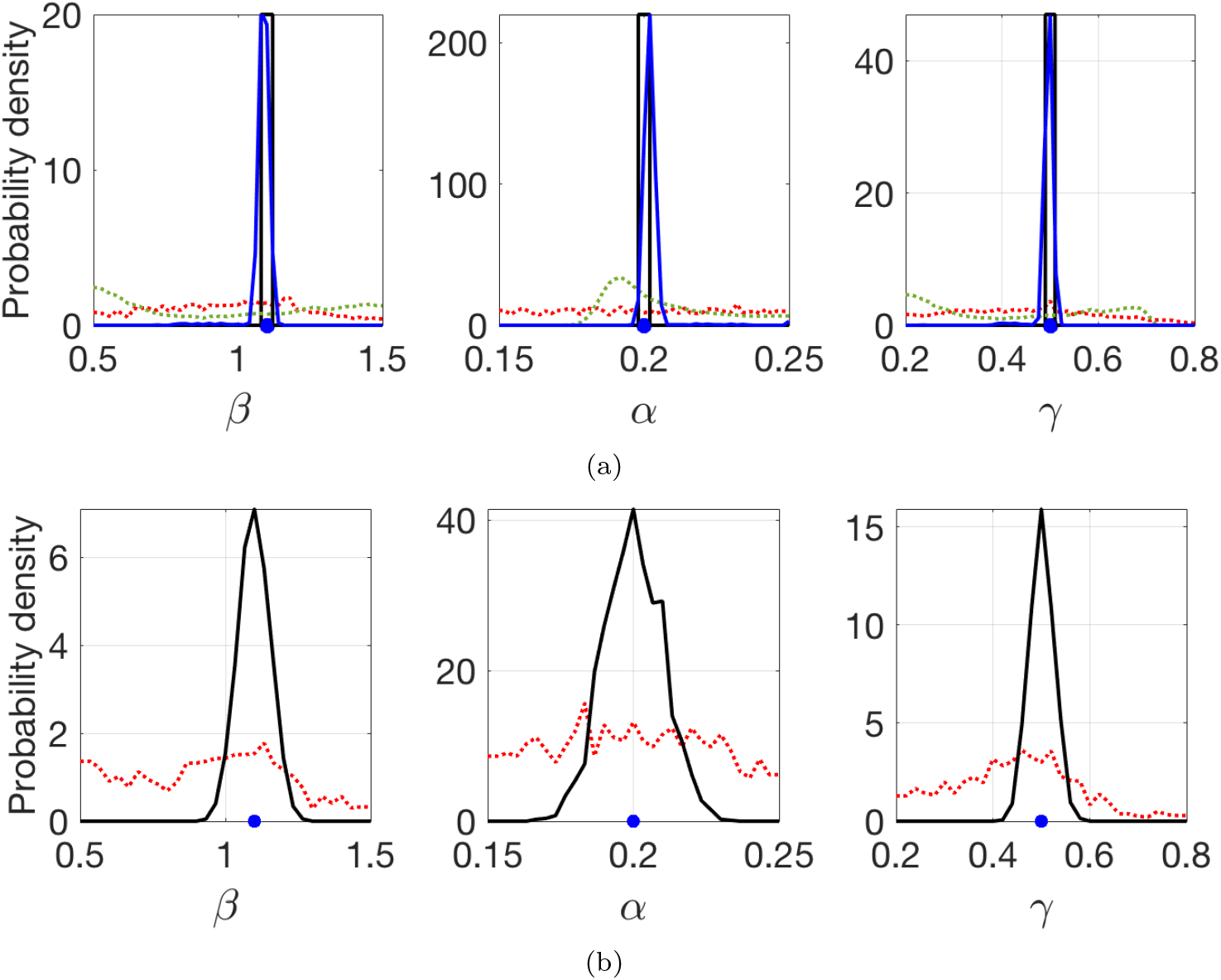
Histograms of estimated parameters from *I*(*t*), collected from the time intervals [0, 50] and [50, 100]. The SEIR model has *β* = 1.5, *α* = 0.2, *γ* = 0.5, and *I*(*t*) was used as input to two different algorithms. Blue dot denotes exact values. (a) Parameters from *I*(*t*) generated by deterministic SEIR. The red (dotted) and black traces use *I*(*t*) from [0, 50] and [50, 100], respectively, by minimizing *L*(*β, α, γ*) from 1000 different trajectories of the deterministic SEIR model. The green (dotted) and blue traces are marginals of the posterior density computed from MCMC using *I*(*t*) from [0, 50] and [50, 100], respectively. (b) Parameters from *I*(*t*) generated by stochastic SEIR. The red and black traces use *I*(*t*) from [0, 50] and [50, 100], respectively as in (a), by minimizing *L*(*β, α, γ*). The MCMC method is not represented in (b), since it would likely be computationally intractable.

The success of this simple approach to parameter estimation on [50, 100] (or the complete interval [0, 100], not shown) is due to the fact that the SEIR model (1) is structurally identifiable from *I*(*t*), as long as the peak of the epidemic can be observed. However, one can see that this approach fails on the outbreak part of the epidemic, as shown by the histogram in red. On the time interval [0, 50], the input *I*(*t*) is not sufficient to constrain the three parameters.

Figure 2(a) also shows a test of a completely different approach to parameter estimation. We applied Markov Chain Monte Carlo (MCMC) to sample the posterior density of the parameters given the observations, namely *P* (*β, α, γ* | Δ*I*_obs_(*t*)) for *t* in the same intervals as above. In the deterministic SEIR (used for the MCMC computation of the posterior) the likelihood *P* (Δ*I*_obs_(*t*) | *β, α, γ*) is a product of Poisson densities which allows easy sampling of the true posterior *P* (*β, α, γ*| Δ*I*_obs_(*t*)). In Fig. 2(a) we show the three marginals of the posterior. We notice similar qualitative behavior for this estimator, namely that the parameters are identifiable from the second half [50, 100] of the epidemic (blue curve), but almost completely unidentifiable from *I*(*t*) during the first half [0, 50] (green curve).

Figure 2(b) returns to minimization of the data assimilation error *L*(*β, α, γ*) as above, but applied to the stochastic SEIR model and using a Poisson-based EnKF. The histogram shows the variation over 1000 different realizations of Poisson noise. For the interval [50, 100], the variation is increased for stochastic SEIR in comparison to the deterministic SEIR, but the estimates are unbiased around the correct parameter settings. For [0, 50], no meaningful estimation occurs.

In summary, for both deterministic and stochastic versions of the SEIR model, both data assimilation-based and MCMC-based algorithms are able to identify the three parameters easily given *I*(*t*) from the time interval [50, 100], and fail on the interval [0, 50]. The intervals [0, 50] and [50, 100] are chosen to be representative of intervals for which identifiability fails and succeeds, respectively. Similarly chosen intervals show the same results, that early in an epidemic, before the peak is reached, there is a structural reason that the parameters will not be identifiable. We address that reason in the next two sections.

## 3. Dynamical compensation in linear models

We will later discuss the fact that during the pre-peak part of the epidemic, the SEIR model is approximately linear, and *E* and *I* are approximately proportional to one another. The goal of this article is to examine how this fact imposes a constraint on our ability to infer parameters from data, in particular from observations of *I*(*t*). The mechanism that causes this is called dynamical compensation. For linear compartmental systems, this phenomenon was reported as early as [2, 3].

### 3.1. Asymptotic behavior of linear models

Consider a linear initial value problem consisting of a vector differential equation 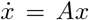, satisfying initial conditions *x*(0) = *x*_0_, where *x* = [*x*_1_, …, *x*_*n*_]. Assume *A* has distinct real eigenvalues. Then solutions are of form

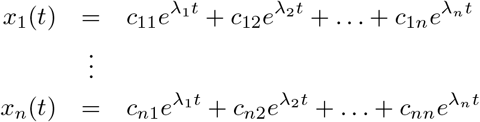

where *λ*_1_ > *λ*_2_ > … > *λ*_*n*_ are the eigenvalues of *A*. Because of the exponential form of the solutions, as *t* moves away from zero, the solutions begin to closely approximate

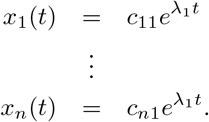

Assuming *c*_11_≠ 0, this means that for each *i, x*_*i*_(*t*) ≈ *c*_*i*_*x*_1_(*t*) for some constant *c*_*i*_.

**Example**. Consider the linear initial value problem

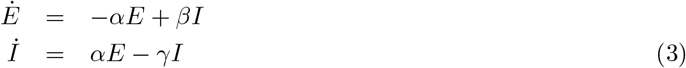

Where we write as 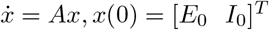 where

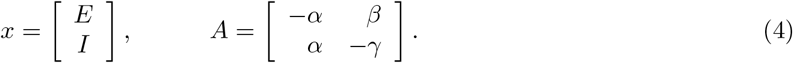

Let *A* = *PDP*^−1^ be the diagonalization, where the columns of *P* are eigenvectors of *A*. The diagonalization exists because *α, β, γ* > 0 implies *A* has distinct real eigenvalues *λ*_1_ > *λ*_2_. The solution is

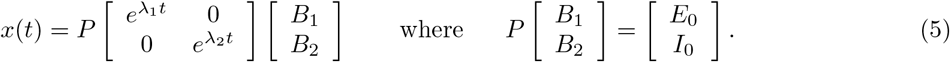

We can consider separate cases, depending on the constants *B*_1_ and *B*_2_. In what we will call the non-generic case, one or both of the *B*_*i*_ is zero. If *B*_1_ = *B*_2_ = 0, the solution is identically zero. If one of the *B*_*i*_ = 0, or equivalently the 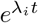 term of the solution is absent, then *I*(*t*) = *cE*(*t*) for some constant *c* and for all *t*.

In the generic case, both *B*_*i*_ ≠ 0, and the solution will be

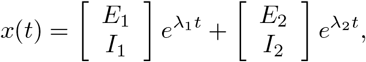

meaning that *I*(*t*) ≈ *cE*(*t*) asymptotically, where *c* = *I*_1_*/E*_1_. Note that in all cases, *I*(*t*) ≈*cE*(*t*) with the approximation improving exponentially in time.

### 3.2. Identifiability in linear systems

A general approach to assessing identifiability in linear systems is suggested in [20]. To search for alternative solutions to (3) with the same output *I*(*t*), but different *E*(*t*) and different parameters (*α*′, *β*′, *γ*′), define the coordinate change *z* = *Sx* for a nonsingular matrix

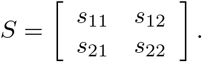

Specifically, we seek an *S* that satisfies

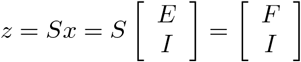

for some *F*. The new variable *z* will reproduce *I*(*t*) as its second entry, using a “dynamically compensating” *F* (*t*) as its first entry, with a different set of parameters, determined below.

This equation is expressible as [0 1]*Sx* = [0 1]*x*. From (5), this constraint is

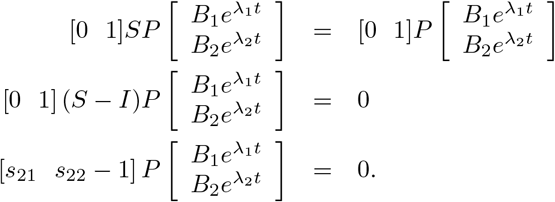

Transposing yields

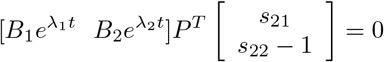

for all *t*. Now we split into two cases, depending on the initial conditions (see (5)).

*Case 1* (generic). In this case, *B*_1_ ≠ 0 and *B*_2_ ≠ 0. Then for two different times *t*_1_, *t*_2_, the rows of the leftmost matrix in

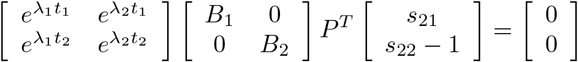

are linearly independent. Since all matrices on the left side are nonsingular, *s*_21_ = 0 and *s*_22_ = 1, and therefore

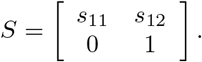

With this change of coordinates, we can consider the alternative system to (3) 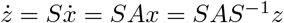 where

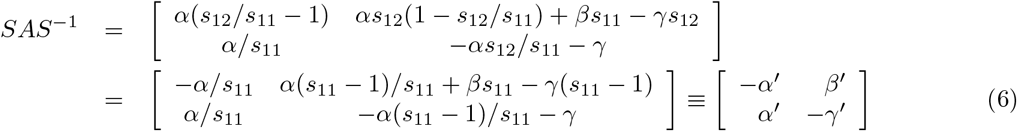

and where we have set *s*_12_ = *s*_11_ −1 to match the desired form (3). This gives a family of alternative solutions of (3) sharing *I*(*t*), but with different parameters and different *E*(*t*), that are indexed by the single parameter *s*_11_. The revised *E*(*t*) is *F* (*t*) = *s*_11_*E*(*t*) + (*s*_11_ − 1)*I*(*t*). These solutions exactly match *I*(*t*) for all *t ≥* 0, and satisfy

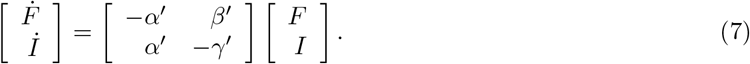

The generic case provides a one-dimensional family of alternative solutions. As promised in [20], these alternative solutions show that in the generic case, the parameters of (3) are unidentifiable from *I*(*t*). That is, on the basis of *I*(*t*) alone, one cannot distinguish between the infinite set of solutions of (7). If our information about the system (3) or its parameters are to be inferred from *I*(*t*), the existence of multiple solutions consistent with the observations of *I*(*t*) will make recovering the parameters effectively impossible.

*Case 2* (non-generic). Now assume that either *B*_1_ or *B*_2_ is zero. Then *I*(*t*) = *cE*(*t*) for all *t*.

The proportionality constant *c* can be calculated from the equations, and depends only on the parameters *α, β, γ*. Keeping the approximation *S* ≈ *N* and substituting *I* = *cE*:

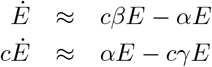

which implies

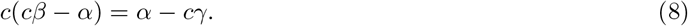

The largest solution *c* of this quadratic equation is real and positive, assuming *α, β, γ* > 0.

**Lemma 1**. *Let α, β, γ* > 0 *and let c* > 0 *be the unique positive solution of the quadratic equation*

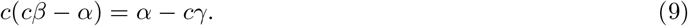

*Define E*(*t*) = *E*_0_e^(c*β* − *α*)*t*^*and I*(*t*) = *cE*(*t*). Let *α*′, *β*′, *γ*′ > 0. *Lie on the surface in* ℝ^3^*defineby*

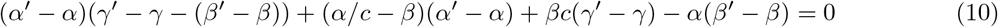

*and define* 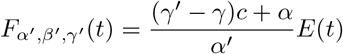. *Then for all α*′, *β*′, *γ*′ *satisfying (10), the set* (*F* = *F* _*α*_′_, *β*_′_, *γ*_′, *I, α*′, *β*′, *γ*′) *satisfy*

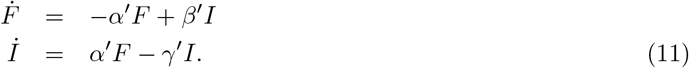

**Proof**. Set *A* = (*γ*′ −*γ*)*c* + *α*, so that 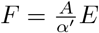.

(i) Note that the right-hand side of the first equation is

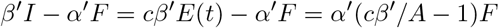

We can calculate

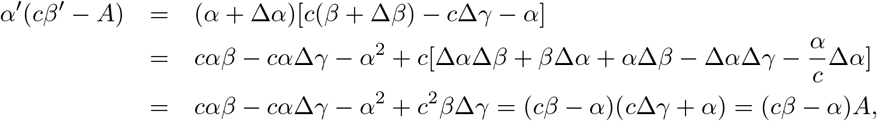

where we have used the notation Δ*α* = *α*′− *α*, Δ*β* = *β*′− *β*, Δ*γ* = *γ*′−*γ*, and used (10) to arrive at the last line. Dividing by *A* recovers *cβ* −*α*. The time derivative of *F* (*t*) is (*cβ* −*α*)*F*, which verifies the first differential equation of (11).

(ii) The right-hand side of the second equation is

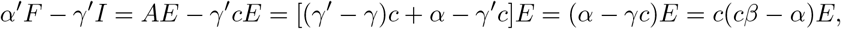

by the quadratic equation (8). This agrees with *İ*, verifying the second differential equation. □

The significance of the lemma is that in Case 2, the equation (10) reveals a two-dimensional family of solutions of (11) with asymptotically identical *I*(*t*), further complicating the identifiability of the parameters. There are substantially more alternative solutions in the non-generic Case 2, a two-dimensional set instead of a one-dimensional set found in Case 1, because we only require that they match *I*(*t*) asymptotically. However, since the asymptotic convergence is exponential, and because infected case counts are often noisiest at the outset of an epidemic, the difference is likely to be insignificant in practical applications. We will return to this observation in the next section, and visualize the comparison in Figure 5.

## 4. Applications to identifiability

In this section, we apply our knowledge of dynamical compensation in linear compartmental models from the last section to the nonlinear SEIR model. We find that in using a linear approximation valid in the pre-peak portion of the epidemic, it is the non-generic case above that turns out to be the most informative on identifiability.

### 4.1. Unidentifiability in pre-peak SEIR

The SEIR model (1) is a coupled set of nonlinear differential equations, but at the beginning of the epidemic, *S*≈ *N*. As the first cases of exposed individuals begin to transition into the infected class, note that the second and third equations approximate a linear system

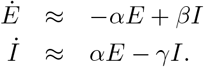

This approximation was exploited in [10] to derive a formula 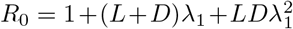 for the reproductive number *R*_0_ = *β/γ* in case *β* is unknown but the latent and infectious periods *L* = 1*/α* and *D* = 1*/γ* and the exponential growth rate *λ*_1_ from (5) can be independently estimated.

According to the previous section, we will observe the asymptotics of the approximately linear dynamics,

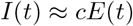

for some *c* as *t* moves away from 0. In fact, this behavior is apparent in Figure 3(b), which is a magnification of panel (a). The trace of *I*(*t*) appears to be a constant proportion of *E*(*t*), and this is confirmed in Figure 3 (c) where the ratio is plotted versus time.

**Figure 3:**
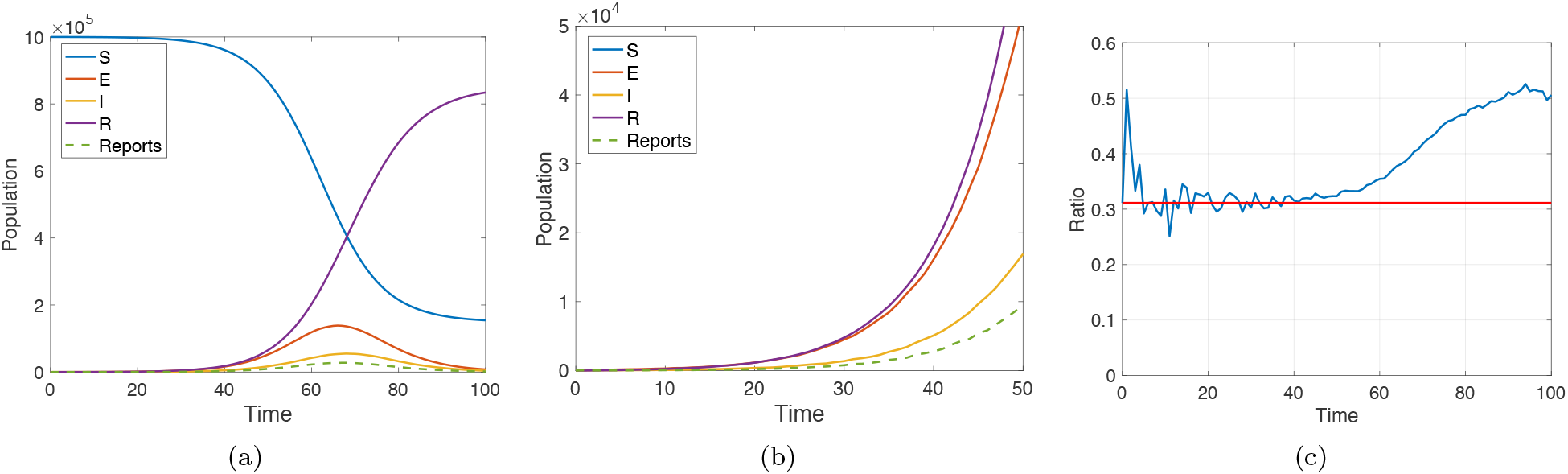
Plot of SEIR populations with parameters *β* = 1.1, *α* = 0.2, *γ* = 0.5. The new cases Δ*I* are denoted by the dashed curve (Reports in the legend). (a) Full plot on [0, 100]. (b) Magnification of (a), restricted to the time interval [0, 50]. (c) The blue curve is a plot of the ratio *I*(*t*)*/E*(*t*). Here *I* ≈ *cE* for the first 50 days, where *c* = 0.31, as calculated from (8).

Figure 4 shows the results of a parameter estimation computation using the data from Figure 3, which sets *β* = 1.1, *α* = 0.2, and *γ* = 0.5. We run data assimilation on the time interval [0, *T*] using only the daily case numbers Δ*I*(*t*) as input, for various choices of *T*. To simplify the situation, we will fix the parameter *γ* = 0.5 to be the exact value, and attempt to estimate *β* and *α*. We accomplish this by minimizing *L*(*β, α*, 0.5) as described in Section 2.2.

**Figure 4:**
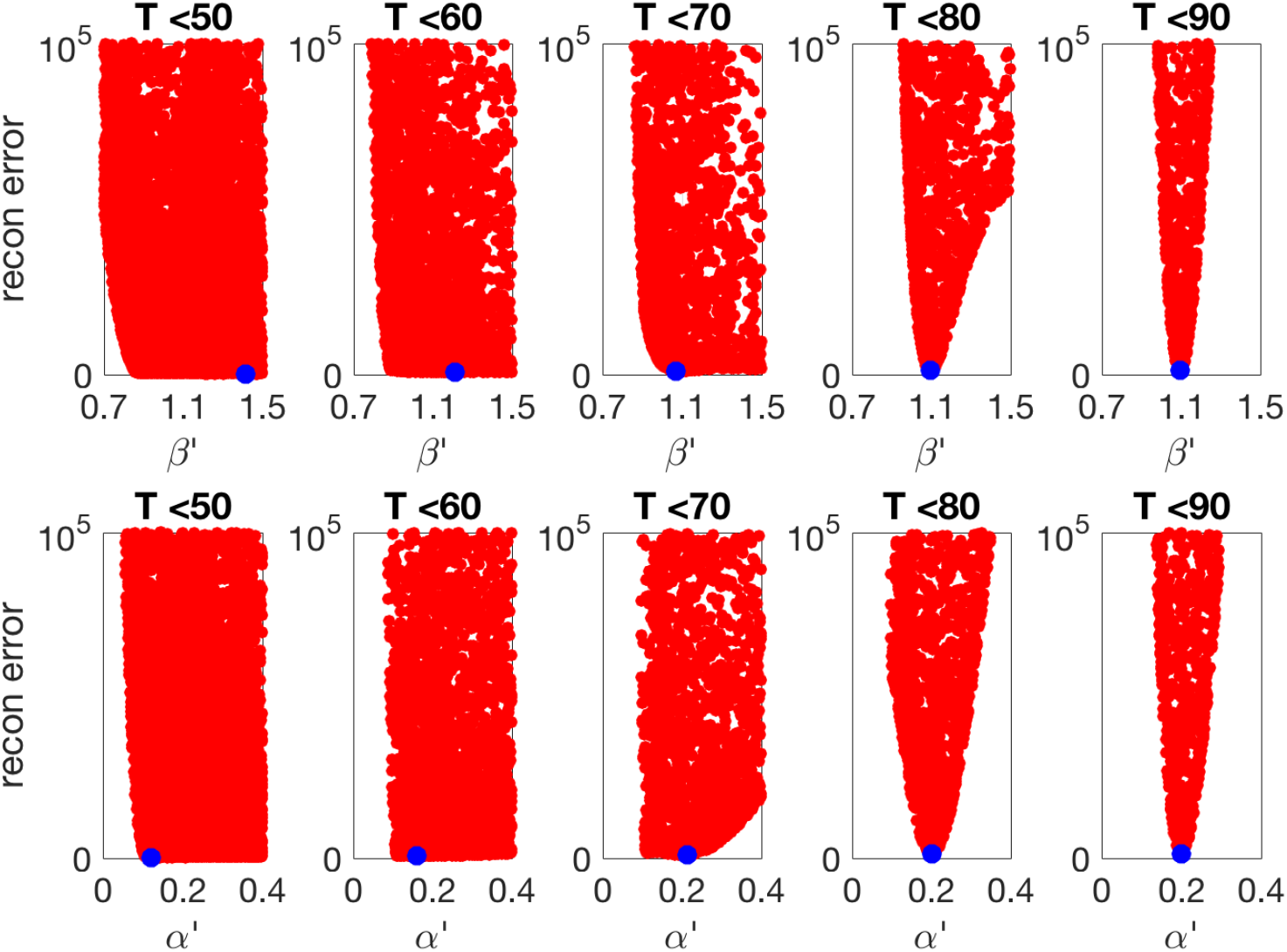
Estimation of parameters by minimization of data assimilation error on the time interval [0, *T*] for various *T*. Each red dot is the value of the sum of squares assimilation error for randomly chosen parameters (*β*′, *α*′), while the exact *γ*′ = *γ* = 0.5 is assumed known. The blue dot represents the calculated minimum. For *T* significantly below 80, the loss function has no well-defined minimum, and the generating parameters (*β* = 1.1, *α* = 0.2) are poorly estimated. For larger *T*, the minimum becomes more pronounced and the parameters can be well estimated.

The function *L*(*β, α*, 0.5), sampled at 10,000 random values, is displayed in Figure 4, projected onto the *β* and *α* axes, respectively, for ease of analysis. For “pre-peak” values of *T*, the parameters *β* and *α* are not well estimated. As *T* increases and approaches the epidemic peak 60 *< T <* 80, the parameter estimates gradually become quite accurate. This corroborates our finding in Figure 2, that parameter estimation fails to isolate correct parameters early in the epidemic.

### 4.2. The unidentifiability manifold

The dynamical compensation results of the previous section explain the phenomenon seen in Figure 4. The unidentifiability manifold, in this case a surface, is plotted in Figure 5. The red dots identify the parameter points (*β*′, *α*′, *γ*′) whose evaluated loss function computed on the time interval [0, 50] is in the lowest 1% of points (among 10,000 random points sampled). The points lie extremely near the unidentifiability surface (10). The wide distribution of the points shows the impossibility of estimating the generating parameter set (*β* = 1.1, *α* = 0.2, *γ* = 0.5) with any accuracy. The color shading on the surface corresponds to reproductive number *R*_0_ = *β*′*/γ*′. We note that *R*_0_ is not significantly constrained by the parameters with minimal loss function.

**Figure 5:**
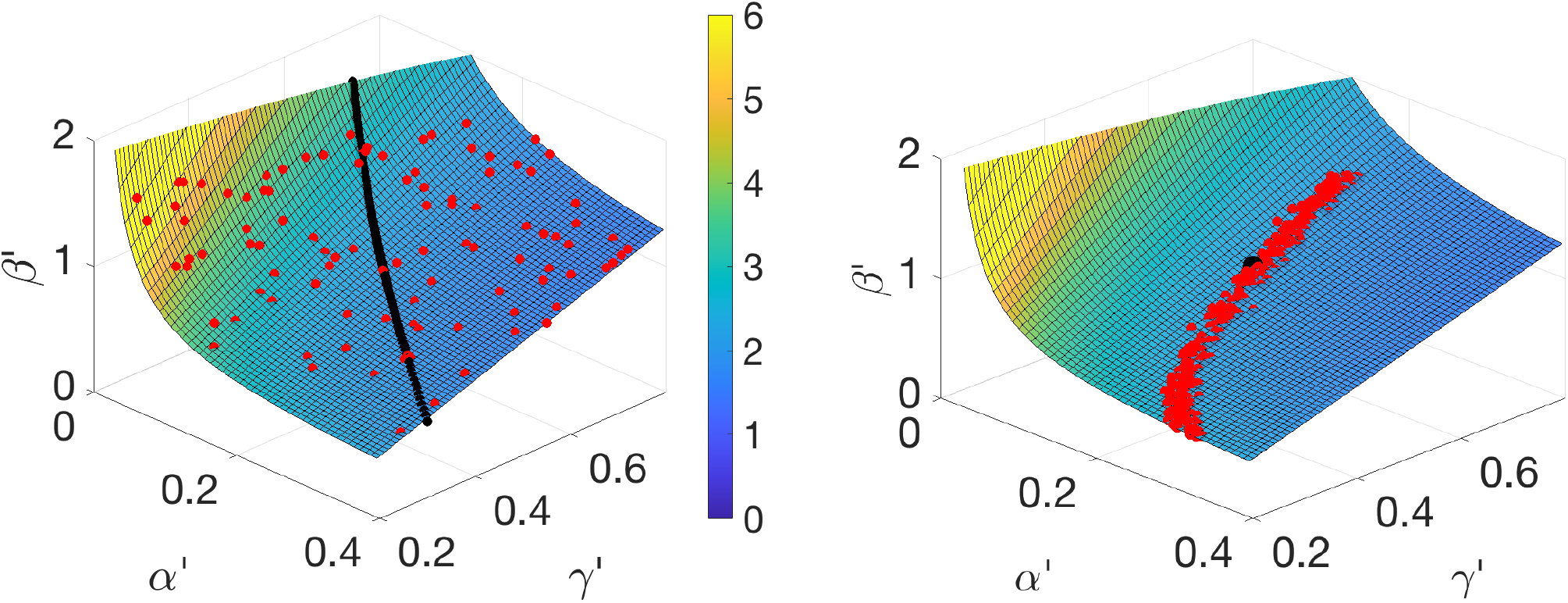
The unidentifiability surface defined by (10). (a) The red plotted points are the parameter values that minimized (landed in the smallest one percent of values) the loss function *L*(*β, α, γ*) from the stochastic nonlinear SEIR model (1) trained on *I*(*t*) from the time interval [0, 50]. They are in remarkable agreement with the quadric surface (10) generated by the “non-generic” solutions. The black curve represents the parameter sets that generate the “generic” solutions from (6). The color represented on the surface corresponds to the computed *R*_0_ = *β*′ */γ* ′. (b) MCMC using *I*(*t*) on [0, 50] from the deterministic version of the nonlinear SEIR (1) to sample the posterior (red dots). They all lie on the surface (10). The true parameters are represented by the black dot.

The MCMC approach introduced in Section 2.2 shows a similar story. In this case, we use observations of the deterministic model (2), and apply MCMC using a single realization of *I*(*t*) in the time interval [0, 50] as observable. The true parameters lie inside the envelope of the posterior, as shown in Fig. 5(b). The Metropolis-Hastings algorithm within MCMC is rejecting thousands of proposals that do not lie on the surface and only accepting those that do.

Since the unidentifiability surface is a two-dimensional set, we conclude that even if one of the parameters is known, the other two are not identifiable – the set of possible parameters will only be reduced to a one-dimensional curve. For example, with fixed *γ*, the data assimilation error on the interval [0, 50] has a poorly-defined minimum as a function of (*β, α*). To illustrate this, fixing *γ* = *γ*′ = 0.5 in in the unidentifiability manifold equation (10) yields the curve *α*^*t*^ = *α* (*β* −*α/c*)*/* (*β*′− *α/c*). This curve is plotted in blue in Figure 6(a). The plotted red points are the one percent of (*β, α*) pairs with smallest values of the loss function. Instead of a localized ball near the true value (*β, α*) = (1.1, 0.2), there is a curve of pairs equally fitting the observed data, which are therefore indistinguishable to the loss function. These pairs form the flat minima of the loss function seen in Figure 4 for times *T* preceding the epidemic peak.

**Figure 6:**
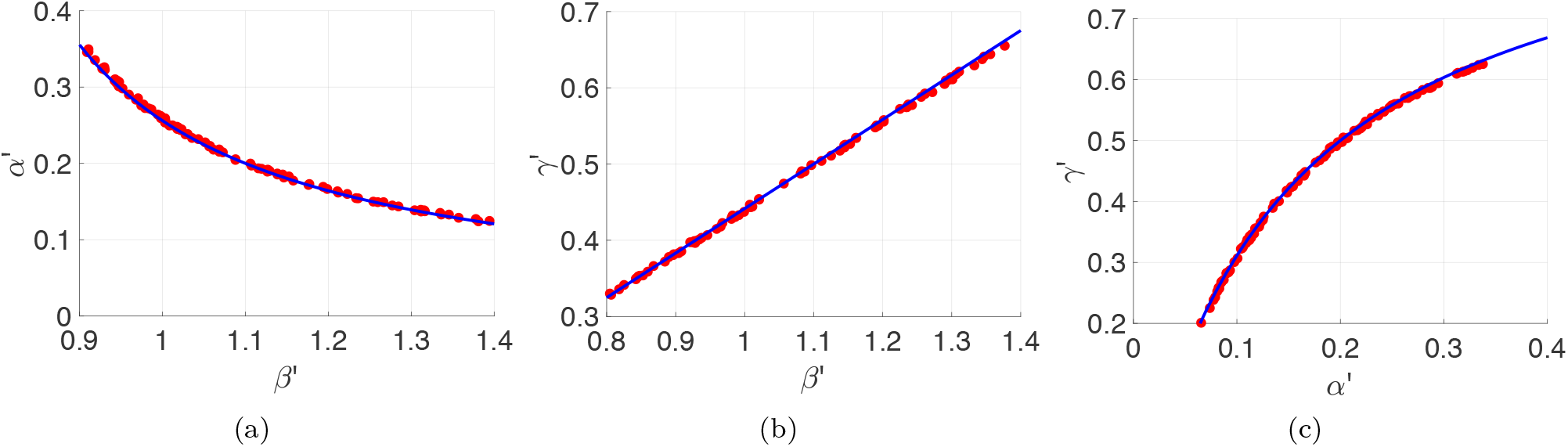
Continua of best parameter sets from the same *I*(*t*). (a) The dots denote the one percent of (*β*′, *α*′) pairs (chosen from 10000 random pairs) with the smallest sum of squares error from data assimilation over the interval [0, 50]. The blue curve is given by equation (10) with *β*^*I*^, *α*′, *c* as in Figure 3, and setting *γ* = *γ*′ = 0.5. (b) The dots are the (*β*′, *γ*′) pairs with smallest assimilation error for fixed *α* = *α*′ = 0.35. Equation (10), plotted as the blue dashed curve, is the line *γ*′ = *γ* + *α* (*β*′ −*I β*)*/* (*cβ*). *I*(c) The dots are the (*α*′, *γ*′) pairs with smallest assimilation error for fixed *β* = *β*′ = 0.7. The red dashed curve is *γ* = *γ* + (*α βc*)*/* (*α*′ + *βc* − *α*) − *α/c* from (10) setting *β* = *β*′ = 0.7.

Similarly, if we fix a different parameter, we see the same phenomena when trying to estimate the other two parameters. For example, fixing *α*′ = *α* = 0.2, the slice through the unidentifiability manifold (10) is *γ*′ = *γ* + *α* (*β*−′ *β*)*/*(*cβ*), a line. Figure 6(b) shows the line in blue, with the near-minimal pairs of the loss function shown as red dots. Finally, fixing *β*′= *β* = 1.1 yields the curve *γ*′ = *γ* + (*β*− *α/c*)*/*(1 + *βc/*(*α*′−*α*)) from the manifold (10), shown in Figure 6(c).

On the other hand, fixing two parameters on the unidentifiability surface implies that the third can be determined. That is, if we have knowledge of the true *α* and *γ*, setting *α*^*t*^ = *α* and *γ*^*t*^ = *γ* in (10) implies that *β*^*t*^ = *β*, so there is a unique solution with those parameter settings. Thus even on the pre-peak interval [0, 50] in the example, if *α* and *γ* are known, then *β* is structurally identifiable from the observations of *I*(*t*).

Of course, there are many other figures of merit that could be minimized to determine the parameters from the observed *I*(*t*), either based on data assimilation errors, maximization of likelihood, or on some other probabilistic measure. However, during the pre-peak part of the epidemic, they will all be susceptible to the alternative solutions that are equally compatible with *I*(*t*), implicit in dynamical compensation.

A perhaps more intuitive view of the unidentifiability surface, if less geometric, is that it is the set of parameters for which the leading eigenvalue *λ*_1_ of the resulting system is equal to the *λ*_1_ (see (5)) of the underlying system that generated *I*(*t*). (In fact, this leads to an alternate derivation of (10).) Thus, if we trust the parameter estimation algorithm to return to us a parameter set that is at least on the unidentifiability surface, then it will have the correct *λ*_1_. Even if the parameters are wrong, this fact can be exploited for uncertainty quantification purposes, as we discuss in the next section.

### 4.3. Uncertainty quantification

The unidentifiability surface (10) is useful for theoretical reasons, to show the impossibility of isolating the original parameter set *p* from the infinity of other systems that approximately share *I*(*t*) during the beginning portion of an epidemic. Next, we suggest that it may be useful in practice for uncertainty quantification.

It turns out to be a helpful fact that the unidentifiability surface generated by an arbitrary parameter set *p* indexes the set of parameter sets that share the observed *I*(*t*). Assume that we use a parameter estimation algorithm with input *I*(*t*), and estimate the parameter set as *p*′, that lies on the surface. The roles of *p* and *p*′ are symmetric, so we can also consider that *p* lies on the unidentifiability surface generated by *p*′. That means we can reverse the roles: switch the primed and unprimed variables in (10), noting that *c* must be replaced by *c*′computed from (9) with unprimed variables replaced with primed variables.

As an illustration, assume the correct parameters are *p* = (*β, α, γ*) = (1.1, 0.2, 0.5) but that a parameter estimation algorithm instead returns, for example, an estimate *p*′ = (*β*′, *α*′, *γ*′) = (0.852, 0.25, 0.4) that lies on the unidentifiability surface. The set *p*′ given here is just for illustration; in this case it was chosen by making an arbitrary choice of *α*^*t*^ and *γ*′, and then computing the corresponding *β*^*t*^ lying on the surface (10). Next, we ignore the origin of *p*′, and consider what we can infer from it. In Figure 7(a), we produce 30 trajectories of the stochastic SEIR by perturbing *p*′ by 10% to new values *p*′′ = (*β*′′, *α*′′, *γ*′′). We have overlaid as a yellow curve the original trajectory that produced *I*(*t*), generated by the parameters *p*. There is a large amount of variability in the 30 trajectories.

**Figure 7:**
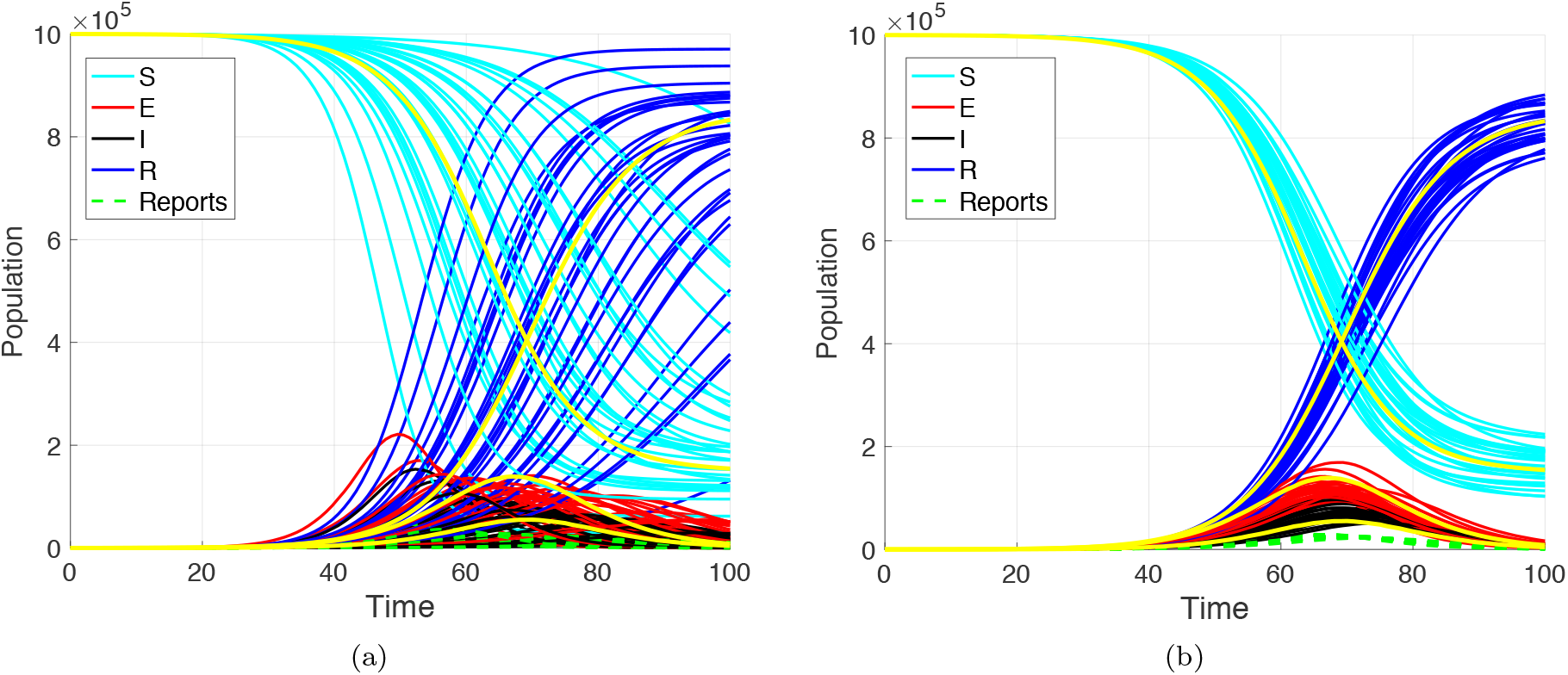
Trajectories of 30 systems with alternative parameter values. (a) Parameter values *p*′′ = (*β*′′, *α*′′, *γ*′′) are chosen by perturbing randomly with 10% Gaussian noise from a fixed *p*′ = (*β*′, *α*′, *γ*′) = (0.852, 0.25, 0.4). The original trajectory with parameter values *p* = (*β, α, γ*) = (1.1, 0.2, 0.5) is traced in yellow. (b) Same as (a), but the *p*′′ are chosen from the surface (10). Specifically, the *p*′′are formed by perturbing (*α*′′, *γ*′′) by 10% and calculating the corresponding *β*′′ lying on the surface.

**Figure 8:**
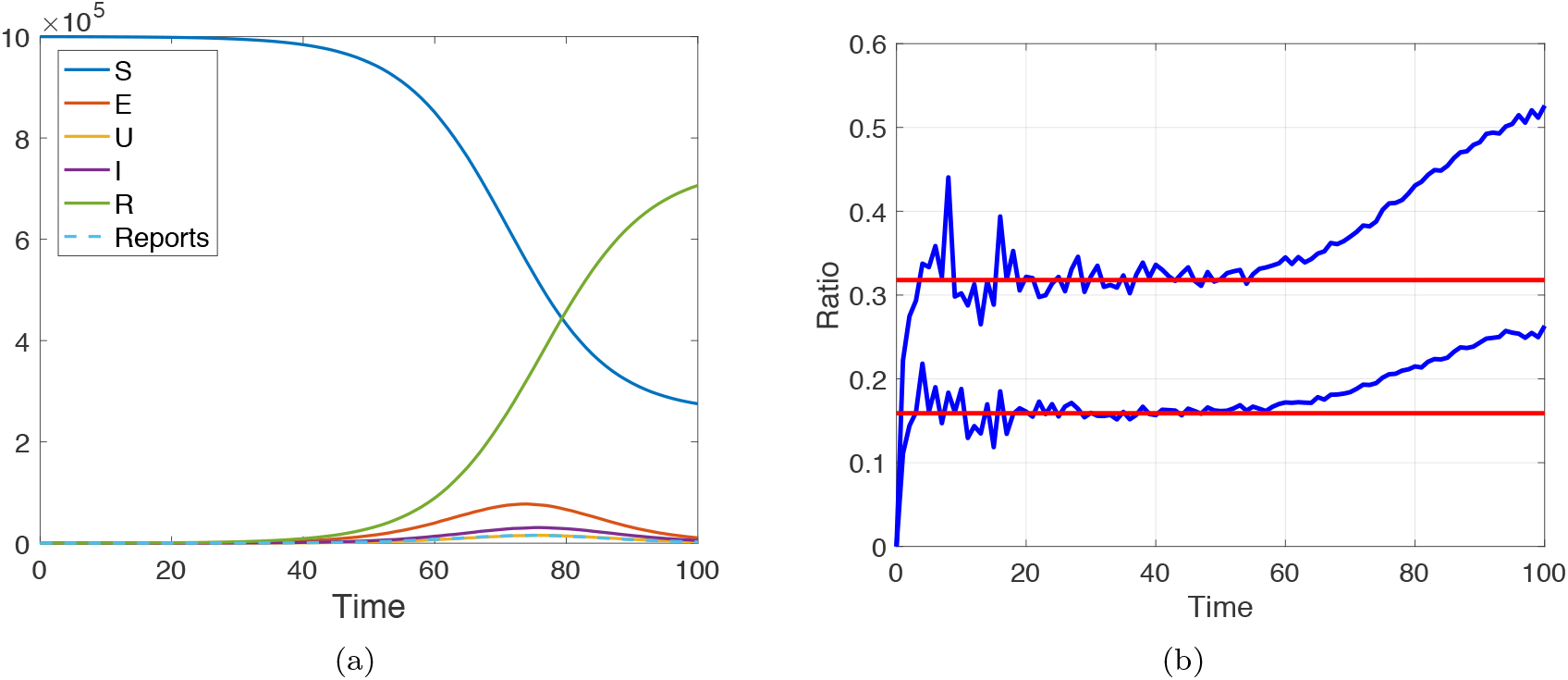
(a) Solution of the stochastic SEUIR equations (13) with initial conditions *S* = 10^6^, *E* = 10^2^, *U* = 0, *I* = 0, *R* = 0. The parameter settings are *β* = 0.9, *z* = 0.3, *w* = 0.2, *d* = 0.5. (b) Ratios *U* (*t*)*/E*(*t*) and *I*(*t*)*/E*(*t*) (blue traces) compared with *b* = 0.16 and *c* = 0.32 calculated from (15), shown in red.

Figure 7(b) shows trajectories of 30 stochastic SEIR systems where we have randomly changed *α*^*t*^ and *γ*^*t*^ by 10% to *α*′′ and *γ*′′, but this time have computed the corresponding *β*′′ that lies on the surface. We reiterate that the surface, being the unidentifiability surface of *p*′, can be computed from *p*′ and is therefore known to us, even if the original *p* is unknown. The ensuing trajectories are much more faithful to the original system, given that they share the leading dynamical eigenvalue *λ*_1_. Thus, even starting with a mildly incorrect parameter set *p*′, by querying nearby points *p*′′ on its unidentifiability surface, we see reasonable facsimiles of the underlying dynamics generated by the original parameters *p*.

Note that there are limitations on how far the incorrect parameters *p*′ can be from the original parameters *p*, in order for the trajectories produced in this way to be representative of the original systems. In particular, the constant *c* in the proportionality *I*(*t*) ≈*cE*(*t*) is in general different for the new system, and so its trajectories will be different. Our informal observation is that if the alternative parameters are within about 20% of the originals, the approximating trajectories may still be useful for uncertainty quantification.

This observation opens up the possibility of using the unidentifiability surface for uncertainty quantification purposes, by studying the spread of nearby solutions as a function of uncertainty in the parameters. If an uncertainty in the estimate can be determined from the algorithm generating the estimate, bootstrapping techniques can be used to move along the surface (10) and quantify the variance of key aspects of the family of nearby trajectories. We leave a more complete analysis of this phenomenon, and its possible utility to forecasting, to future investigation.

## 5. Identifiability in other SEIR-like models

The same identifiability problems are likely to occur in models similar to SEIR. We describe the details for one such example that was proposed recently in [14].

### 5.1. The SEUIR model

In [14], the model was used to represent populations in a specific city, and included extra external inputs from other cities. The underlying SEIR-style model is

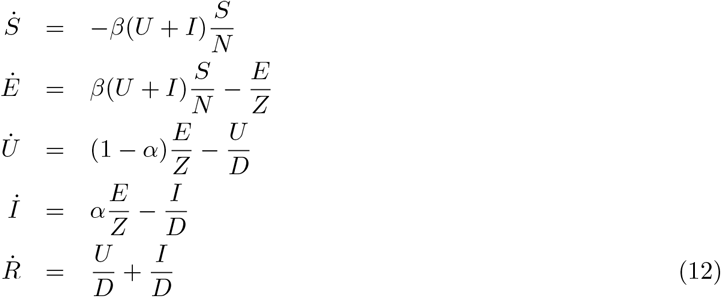

with constant total population *N* = *S* + *E* + *U* + *I* + *R*, where 0 *< α <* 1. The new variable *U* represents unreported infected cases, while *I* is reserved for reported infected cases. As for SEIR, we will consider *I*(*t*) as the observable variable.

For simplicity, we rewrite the parameters as *z* = 1*/Z, d* = 1*/D, w* = *α/Z* to arrive at the equivalent but more user-friendly system

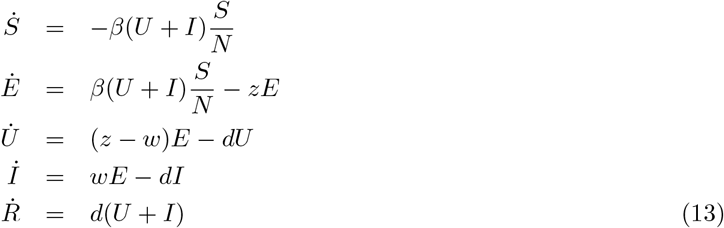

where *N* = *S* + *E* + *U* + *I* + *R*, with parameters *β, z, w* and *d*, 0 *< w < z*, which we call the SEUIR model.

### 5.2. Unidentifiability in SEUIR

Again consider the pre-peak portion of the epidemic, where *S*≈ *N*. Then there is an approximating linear system

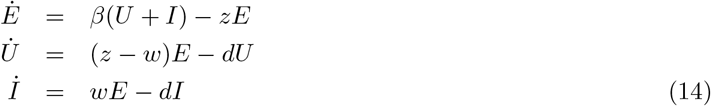

which will exhibit dynamical compensation. Given our experience with SEIR, we consider solutions of (14) where *E, U* and *I* are proportional, say *U* (*t*) = *bE*(*t*) and *I*(*t*) = *cE*(*t*). One checks that if *E*(*t*), *U* (*t*), *I*(*t*) are such solutions, then *E*(*t*) = *E*_0_*e*^[*β*(*b*+*c*)−*z*]*t*^ where

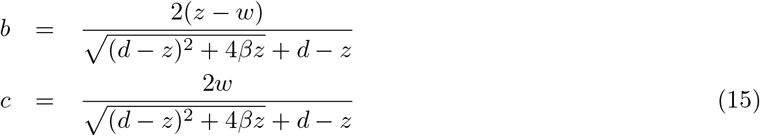

It will be convenient in proving the lemma below to note the identities

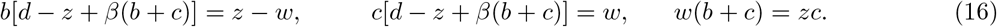

**Lemma 2**. *Let β, z, w, d* > 0 *and E*(*t*), *U* (*t*), *I*(*t*) *be solutions of (14). Let β*′, *z*′, *w*′, *d*′> 0 *and consider the functions*

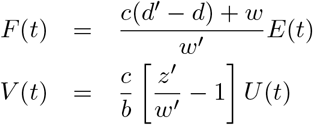

*where b and c are defined in (15). Assume that β*′, *z*′*and d*′ *lie on the surface defined by*

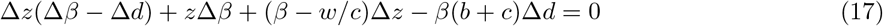

*where we denote* Δ*β* = *β*^*t*^ *β*, Δ*z* = *z*′ *z*, Δ*d* = *d*′*d*.

*Then for all β*′, *z*′, *d*′ > 0 *satisfying (17) and any* 0 *< w*′ *< z*′, *the set*

(*F* = *F*_*β*_, *z, w, d, V* = *V*_*β*_, *z, w, d, I, β*′, *z*′, *w*′, *d*′) *satisfies*

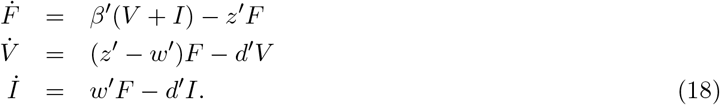

**Proof**. The left-hand side of the first equation is

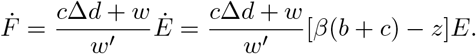

The right-hand side is

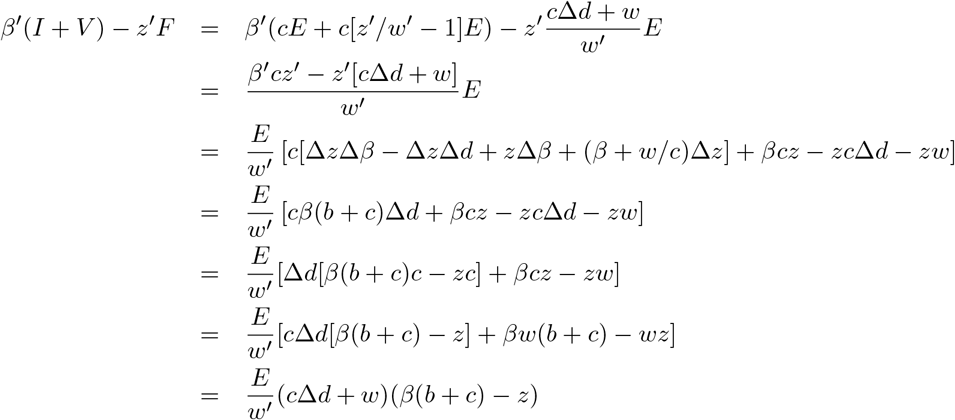

where we used the unidentifiability surface equation (17), and used the identity *w*(*b* + *c*) = *zc* from (16) in the penultimate line. This matches the left-hand side.

The second and third equations use only the definitions of *F* and *W*. For the second equation,

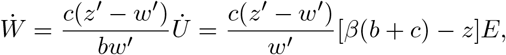

and the right-hand side is

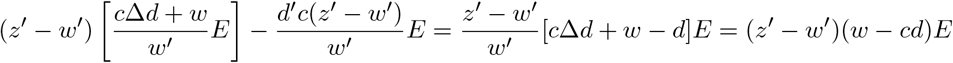

which agrees with the left side by (16). The left side of the third equation is

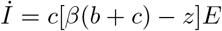

which matches the right side

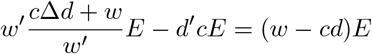

by (16). □

Figure 9 shows a plot of the unidentifiability surface in ℝ^3^, along with a plot of the one percent of random parameter sets (*β, z, d*) that have the lowest loss function values from the nonlinear SEUIR model, using *I*(*t*) as input, on the pre-peak interval [0, 50]. The generating parameters were *β* = 0.9, *z* = 0.3, *w* = 0.2, and *d* = 0.5. These parameter sets will be practically indistinguishable when attempting parameter estimation with *I*(*t*) only over this interval. Here the *w* parameter value has been fixed at the generating value *w* = 0.2.

**Figure 9:**
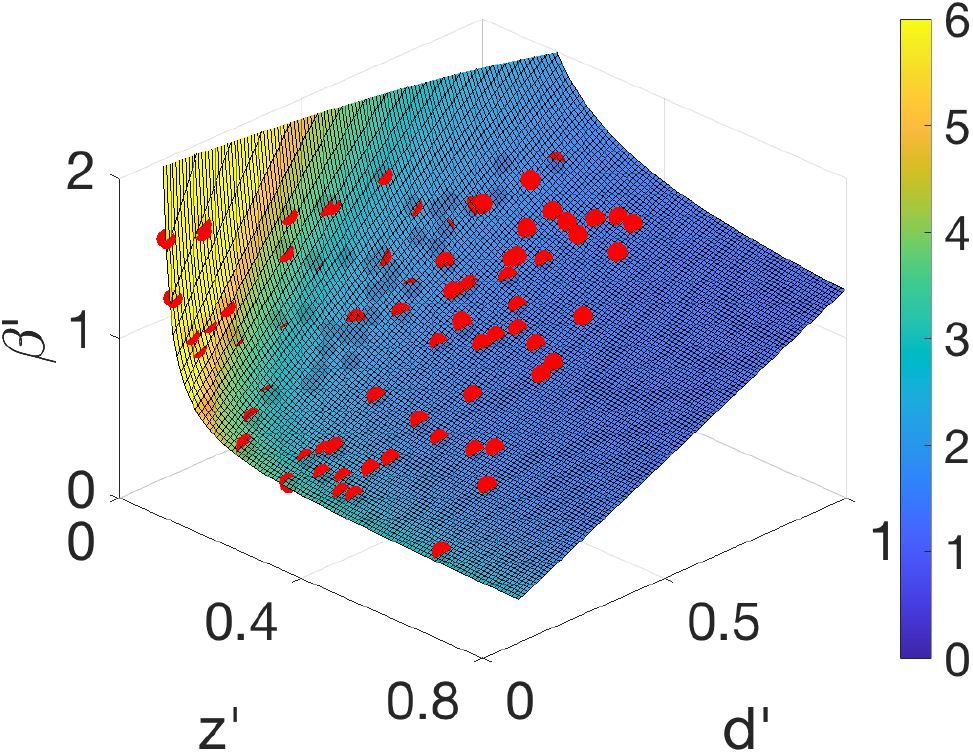
The unidentifiability surface defined by (17). The red plotted points are the parameter values that minimized (landed in the smallest one percent of values) the loss function *L*(*β*′, *z*′, *d*′) from the nonlinear SEUIR model (13), where *w*′ = *w* = 0.2 was assumed known. The parameters generating the input *I*(*t*) were (*β, z, w, d*) = (0.9, 0.3, 0.2, 0.5). The input *I*(*t*) was used from the pre-peak time interval [0, 50]. The surface is colored corresponding to *R*_0_.

Figure 10 shows the results of repeating the sampling of the loss function while fixing *w* = 0.2 and a second parameter. For example, in Figure 10(a) the best one percent of parameter sets (*β, z*) are plotted as dots, along with the relation (17) with Δ*d* set to 0. The relation, plotted as a curve, is *z*′ = *z*(*β*^*t*^ −*w/c*)*/*(*β*^*t*^− *w/c*), and matches the data accurately. In Figure 10(b), the parameter *z*′ = *z* = 0.3, and Δ*z* = 0 in (17) gives a line *d*′ = *d* + *z*(*β*′− *β*)*/*(*β*(*b* + *c*)). In Figure 10(c) with *β*′ = *β* = 0.9, the curve is *d*^*t*^ = *d* + (*β* −*w/c*)(*z*′− *z*)*/*(*z*′− *z* + *β*(*b* + *c*).

**Figure 10:**
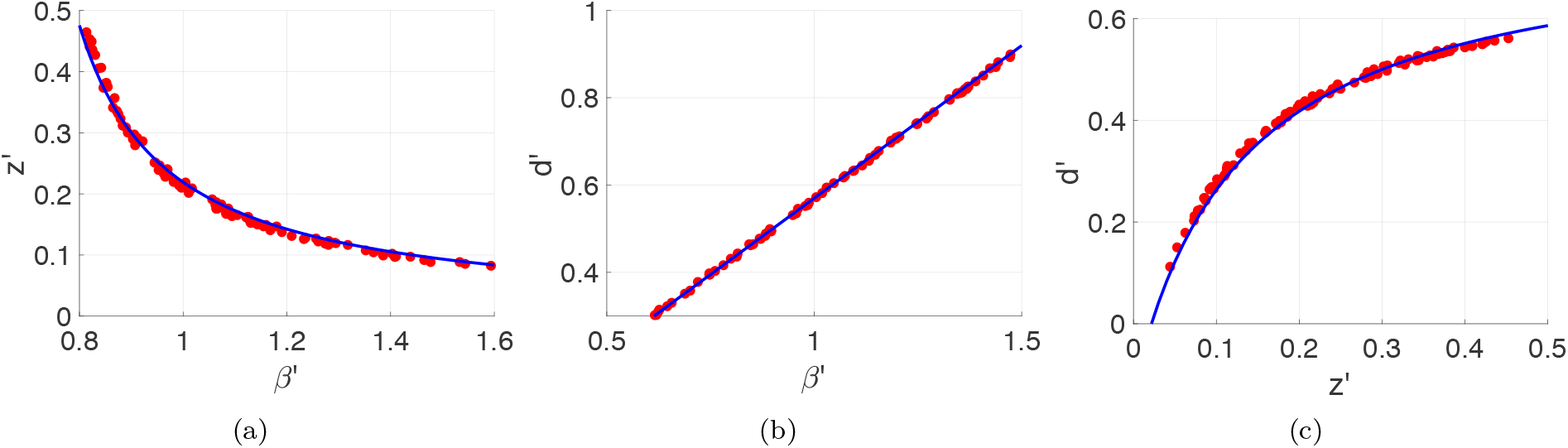
Continuous families of best parameter sets that share the same *I*(*t*). The dots denote the one percent of pairs (chosen from 10000 random pairs) with the smallest sum of squares error *L* from data assimilation over the interval [0, 50]. The solid curve is (17) with Δ*w* = 0 and (a) Δ*d* = 0, (b) Δ*z* = 0, (c) Δ*β* = 0.

The identifiability problem with SEUIR is arguably worse than for SEIR, since a glance at the unidentifiability relation (17) shows no Δ*w* term. Thus the multiple solutions of Lemma 2 exist for any value of *w*′ *< z*′. These solutions have (*β*′, *z*′, *d*′) independent of *w*′, while having adjusted *F* (*t*) and *W* (*t*) that do depend on *w*^*t*^. This results in an added dimension of unidentifiable parameters. In other words, Figures 9 and 10 can be reproduced identically if *w*′ is fixed at an inaccurate value *w*′ = *w*. This means that the actual unidentifiability set is a two-dimensional set in ℝ^4^ of points (*β*′, *z*′, *w*′, *d*′) satisfying (17) and all *w*′ such that 0 *< w*′ *< z*′.

A final comment about the SEUIR model (13) is that one can introduce the new variable *Y* = *U* + *I* and arrive at the equivalent SEIR system

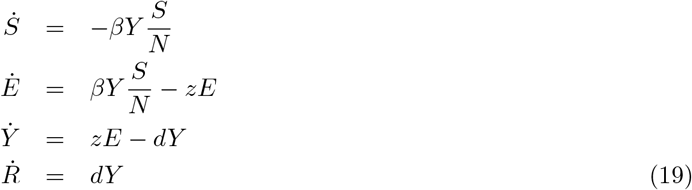

where *N* = *S* + *E* + *Y* + *R*. This may explain the disappearance of the parameter *w*′ in the unidentifiability surface equation (17). However, under the model (13), the assumption is that *I*(*t*) is observed, not *Y* (*t*).

## 6. Discussion

In common epidemic models, practical identifiability from the infected cases variable *I*(*t*) depends strongly on what portion of the population trajectory is observed. In the pre-peak interval, when *S*(*t*) ≈*N*, the linear approximation to the full model admits an infinity of solutions with the same *I*(*t*) by adjusting the unobserved population variables to compensate, a property known as dynamical compensation. The combinations of parameters that allow for this compensation are given by (10) and (17) in Lemmas 1 and 2, in what we call the unidentifiability surface, or unidentifiability manifold. The multiple solutions that coexist in this scenario will defeat any parameter estimation method that relies on observing only *I*(*t*) to find the complete set of parameters. Since the unidentifiability manifold is two-dimensional, at least two more independent pieces of information are necessary to isolate any of the parameters. This also applies to most combinations of the parameters, such as the reproductive rate *R*_0_. These obstructions to identifiability disappear if the entire time history, including the peak of the epidemic, can be observed.

We have shown these identifiability obstructions exist for the popular SEIR model and another more recent model. It is likely that any other closely-related version of SEIR, including compartmental models such as SEIRS, SIRD, etc. will harbor similar obstructions, due to the same phenomenon.

It is notable that the unidentifiability surfaces found for both models are codimension one in parameter space. We conclude that if all but one of the parameters is known a priori, then that last parameter can be determined from an estimation process like the minimization technique used here, even during the pre-peak portion of the epidemic. We have also proposed that knowledge of the unidentifiability surface may be crucial for the development of practical uncertainty quantification for parameter estimates, although pursuit of that direction is beyond the scope of this article.

## Data Availability

No data was used in the preparation of this manuscript.

## Acknowledgements

This work was supported by a National Institutes of Health Director’s Transformative Award (1R01AI145057) and the National Science Foundation (DMS-1723175 and DMS-1854204). MMN was supported in part by the Covid Virus Seed Fund (award 8013) from the Institute for Computational and Data Sciences and the Huck Institute at the Pennsylvania State University.

## Notes

### Competing Interest Statement

The authors have declared no competing interest.

### Funding Statement

This work was supported by a National Institutes of Health Directors Transformative Award (1R01AI145057) and the National Science Foundation (DMS-1723175 and DMS-1854204). MMN was supported in part by the Covid Virus Seed Fund (award 8013) from the Institute for Computational and Data Sciences and the Huck Institute at the Pennsylvania State University.

### Author Declarations

No IRB oversight was necessary in the preparation of this work.

